# Weight Loss With Microdose Tirzepatide: Real-World Outcomes in Patients Initiating Treatment Below 2.5 mg Weekly

**DOI:** 10.64898/2026.09.15.26363172

**Authors:** Jay Hastings, Jayden Lee

## Abstract

**Background:** Tirzepatide is conventionally initiated at 2.5 mg weekly and titrated to approved maintenance doses. Evidence describing weight change at injectable starting doses below 2.5 mg/week is limited.

**Methods:** We conducted a retrospective, single-organization observational study of adults documented as new GLP-1 starts, with baseline body mass index (BMI) ≥27 kg/m^2^, who initiated compounded injectable tirzepatide at a microdose of 1 or 2 mg/week (below the standard 2.5 mg/week starting dose). Source records were reviewed for baseline validity, prior treatment, weight consistency, and treatment exposure. The main descriptive analysis selected the latest eligible measurement per patient within observation windows of 14–35, 36–59, and 60–89 days of adjudicated treatment exposure, excluding observations after escalation above 2 mg/week. All 93 eligible measurements were included in a supporting random-intercept model. Analyses were retrospective and hypothesis-generating; no formal sample-size calculation was performed.

**Results:** Sixty patients contributed 93 eligible follow-up measurements, of which 84 were selected for patient-window summaries. Mean age was 47.3 years, 81.7% were female, and mean baseline BMI was 33.3 kg/m^2^. Mean body-weight loss was 2.18% (95% CI 1.61–2.76; n=57) in the 14–35-day window at a median 21 days, 3.58% (95% CI 2.43–4.72; n=19) in the 36–59-day window at a median 48 days, and 6.10% (95% CI 4.60–7.60; n=8) in the 60–89-day window at a median 65.5 days. The adjusted model estimated 2.16 percentage points greater loss per 30 days of treatment exposure (95% CI 1.69–2.63).

**Conclusions:** Weight loss was observed within the 14–35-, 36–59-, and 60–89-day observation windows among evaluable adults initiating compounded injectable tirzepatide at 1–2 mg/week. These uncontrolled findings are hypothesis-generating; the 36–59- and 60–89-day estimates are particularly limited by sparse follow-up.

**Research in context:** *Evidence before this study:* Randomized trials demonstrate substantial long-term weight reduction with tirzepatide 5–15 mg/week after initiation at 2.5 mg/week. Real-world studies also support effectiveness, but published evidence specifically evaluating tirzepatide initiation below 2.5 mg/week remains sparse. We did not identify a randomized comparison of 1–2 mg/week initiation versus the labeled schedule.

*Added value of this study:* This study applies source-record validation and treatment-start adjudication to 93 follow-up weight measurements from 60 documented new starts. It describes weight change during treatment at 1–2 mg/week and distinguishes individual measurements from patient-window summaries.

*Implications:* The findings support prospective comparative evaluation of lower-dose tirzepatide initiation with standardized weight and adverse-event measurement.

## Introduction

Tirzepatide is a dual glucose-dependent insulinotropic polypeptide and glucagon-like peptide-1 receptor agonist approved for chronic weight management in eligible adults. The U.S. prescribing information recommends 2.5 mg subcutaneously once weekly for four weeks, followed by escalation; 2.5 mg is an initiation dose rather than an approved maintenance dose.^1,2^ Randomized trials using maintenance doses of 5–15 mg/week have demonstrated substantial and sustained weight reduction.^1,10^

Use outside trial protocols is more heterogeneous. Patients and clinicians may consider lower initial doses because of tolerability, cost, or individualized goals. Real-world implementation of these lower initial doses often occurs within formalized programs, such as the PlexusDx Microdose GLP-1 Protocol (https://plexusdx.com/products/microdose-glp1-protocol). However, compounded products are not FDA-approved, and regulators have raised concerns about dosing errors and variable presentation.^8^ Outcomes below the labeled 2.5 mg weekly starting dose warrant investigation without assuming comparative benefits.

We examined weight change among adults initiating compounded injectable tirzepatide below 2.5 mg/week. Throughout this study, we use the term “microdose” strictly to define initiation and treatment at doses below the standard 2.5 mg/week labeled starting dose. The analysis was restricted to observations while documented treatment remained at 1–2 mg/week; measurements after escalation above 2 mg/week were not attributed to this microdose regimen.

## Methods

### Study design and setting

This retrospective observational study used routine-care records from the PlexusDx Telehealth Weight-Management Program (https://plexusdx.com/pages/weight-management-protocols). The available intake/refill export spanned May 29–September 2, 2026. In the final analytic cohort, adjudicated treatment-start dates ranged from June 4 to August 17, 2026, and analyzed follow-up weight dates ranged from June 22 to September 1, 2026. These source-record, treatment-initiation, and outcome dates are distinct. The data freeze was September 2, 2026. The report is organized using STROBE guidance.^7^

### Participants

Adult candidate cases were prescribed injectable tirzepatide at <2.5 mg/week; follow-up availability was assessed subsequently rather than required to enter the candidate pool. The primary evaluable cohort required documented new-start status without identified prior/recent GLP-1 treatment, an acceptable baseline height and weight, BMI ≥27 kg/m^2^, and at least one valid follow-up after ≥14 days of adjudicated continuous treatment exposure at ≤2 mg/week. Cases classified as prior exposure, transfer, restart, or uncertain new start were excluded, as were unsuitable routes or medications and unreliable baselines.

New-start classification reflects the available clinical documentation and does not independently establish lifetime absence of exposure.

### Data validation and exposure adjudication

A cohort-wide screen flagged large or internally inconsistent weight changes, duplicate submissions, discrepancies between absolute weight and contemporaneous reported loss, and source conflicts. Flagged observations underwent case-level review using forms, provider/support notes, messages, prescriptions, payments, orders, and shipping records. No corrected weight was inferred by arithmetic. Unresolved conflicting observations were excluded from the primary analysis and retained in the audit records. Cleaning and analysis decisions evolved during retrospective review, during which provisional outcomes were inspected; they were not a prospectively registered, outcome-blinded protocol.

Treatment start was assigned hierarchically from explicit first-dose evidence, documented receipt/delivery, shipment/fulfillment evidence, other evidence of medication availability, or a provider-decision proxy when no better source existed. Elapsed treatment-exposure days were calculated as weight date minus adjudicated start date. This is an estimate of elapsed time, not proof of daily adherence or first administration. Affected observations after a documented ≥2-week interruption, medication switch, route change, material dosing deviation, or escalation above 2 mg/week were excluded from the primary analysis; valid pre-event observations could remain. Underlying records were not deleted.

### Outcome measures and observation windows

The main descriptive outcome was percentage body-weight loss, calculated as (baseline weight − follow-up weight)/baseline weight ×100; positive values denote loss. Additional outcomes were absolute pounds lost and proportions reaching ≥3% and ≥5% loss. Observation windows were defined by adjudicated treatment exposure: early, 14–35 days; intermediate, 36–59 days; and later, 60–89 days, inclusive. Exact ranges are used below. These labels describe follow-up timing, not biological stages or evidence strength.

Patient-reported tolerability was summarized separately at the earliest eligible frozen follow-up with a date-matched exported response. The export contained a Side Effects field with Yes/No responses and symptom text, and a separate overall-feeling field. No formal adverse-event severity grading or causality adjudication was performed.

### Statistical analysis

The latest eligible measurement per patient within each observation window was selected. A patient could contribute to multiple windows but only once within a window. Means are reported with two-sided 95% t-distribution confidence intervals; medians and responder fractions are descriptive. Paired t tests summarized within-patient changes. A Gaussian random-intercept model used all 93 eligible measurements, with continuous elapsed treatment-exposure days, baseline BMI, age, and sex as fixed effects and a patient-specific random intercept; it was fitted by maximum likelihood. No formal sample-size calculation was performed: sample size was determined by available eligible records. All analyses are exploratory and hypothesis-generating, including the 14–35-day description; no sample-size threshold distinguishes confirmatory from exploratory results. Confidence intervals and P values are nominal and unadjusted for multiple analyses. Missing weights were not imputed. Python, pandas, NumPy, SciPy, and a directly implemented maximum-likelihood model were used.

Sensitivity analyses selected the first eligible measurement per patient-window and, separately, shifted all estimated adjudicated starts by three or seven additional days while leaving the one verified start unchanged. Exposure days and window membership were recomputed, measurements below 14 days were excluded, and latest-per-window selection was reapplied. These are hypothetical timing scenarios conducted on the frozen data, not verified changes to treatment dates.

## Results

### Cohort selection and characteristics

The source audit identified 274 adult candidate cases. Excluding 53 with prior exposure, transfer, restart, or uncertain status and one unusable baseline left 220 documented new starts with acceptable baselines; 127 had valid audited follow-up, of whom 73 had BMI ≥27 kg/m^2^. Exposure adjudication and the ≥14-day, continuous 1–2 mg/week restriction left 60 patients and 93 eligible follow-up measurements (Figure 1).

**Figure 1.**
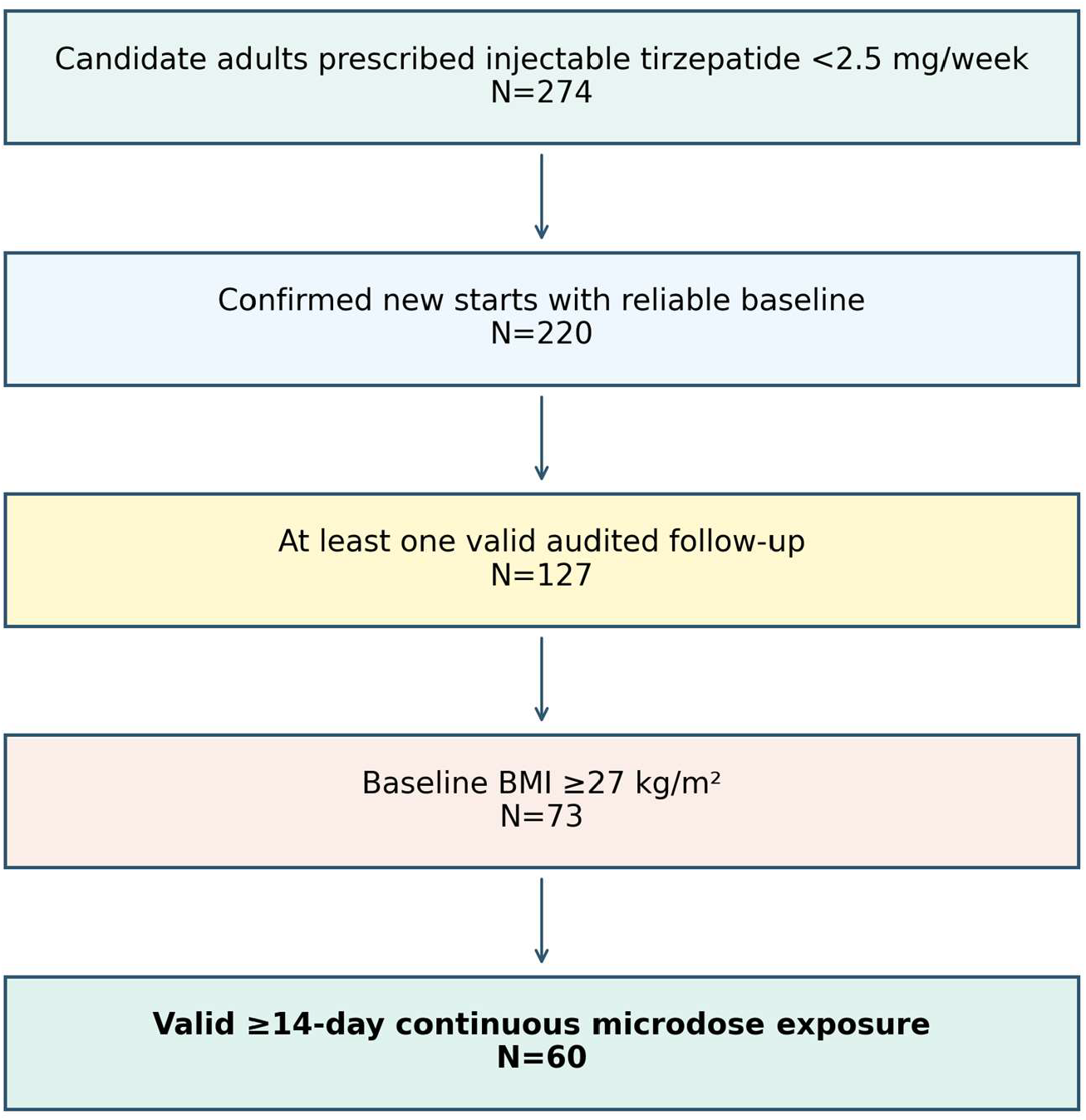
Cohort selection. Candidate entry did not require follow-up. Of 73 BMI-eligible patients, 60 contributed at least one eligible observation after treatment-exposure restrictions; reasons and timing restrictions were applied at patient or observation level as appropriate.

Among the 73 assessed for exposure, two starts were verified, 70 estimated, and one unestablished; among the final 60, one was verified and 59 estimated. In the 72 with assignable starts, provider approval preceded adjudicated start by a median of six days. No eligible observation reached ≥90 days.

### Weight change by observation window

Mean body-weight loss was 2.18% (95% CI 1.61–2.76) among 57 patients in the 14–35-day observation window, 3.58% (95% CI 2.43–4.72) among 19 in the 36–59-day window, and 6.10% (95% CI 4.60–7.60) among eight in the 60–89-day window (Table 2; Figure 2). The corresponding 63, 21, and nine eligible measurements yielded 57, 19, and eight selected patient-window records. Thus, 93 measurements yielded 84 patient-window summaries; six, two, and one additional same-window measurements were omitted only from those summaries and retained in the mixed model.

**Table 1.** Baseline characteristics of the final analytic cohort.

| Characteristic | Value |
| --- | --- |
| Patients, n | 60 |
| Observations | 93 |
| Age, mean (SD), y | 47.3 (11.0) |
| Female, n (%) | 49 (81.7%) |
| Baseline BMI, mean (SD), kg/m<sup>2</sup> | 33.3 (6.1) |
| Baseline BMI, median (IQR), kg/m<sup>2</sup> | 31.4 (28.7–36.1) |
| Baseline weight, mean (SD), lb | 205.7 (47.6) |
| Initiated 1 mg/week, n (%) | 53 (88.3%) |
| Initiated 2 mg/week, n (%) | 7 (11.7%) |

**Table 2.** Weight outcomes and reconciliation of measurements with patient-window summaries.

| Measure | 14–35 days | 36–59 days | 60–89 days |
| --- | --- | --- | --- |
| Eligible follow-up measurements | 63 | 21 | 9 |
| Patients / selected patient-window records | 57 | 19 | 8 |
| Additional same-window measurements | 6 | 2 | 1 |
| Treatment exposure, median days | 21 | 48 | 65.5 |
| Weight loss, mean % (95% CI) | 2.18 (1.61–2.76) | 3.58 (2.43–4.72) | 6.10 (4.60–7.60) |
| Weight loss, median % | 1.96 | 3.59 | 5.88 |
| Absolute weight loss, mean lb | 4.4 | 6.9 | 12.2 |
| ≥3% weight loss, n/N (%) | 12/57 (21.1) | 11/19 (57.9) | 8/8 (100.0) |
| ≥5% weight loss, n/N (%) | 5/57 (8.8) | 5/19 (26.3) | 6/8 (75.0) |

Each patient contributes at most one (latest) measurement per window; the same patient may appear in more than one column. The 84 selected patient-window records represent 60 people, not 84 distinct participants. All estimates are descriptive; the n=19 and n=8 windows warrant particular caution. CI, confidence interval.

**Figure 2.**
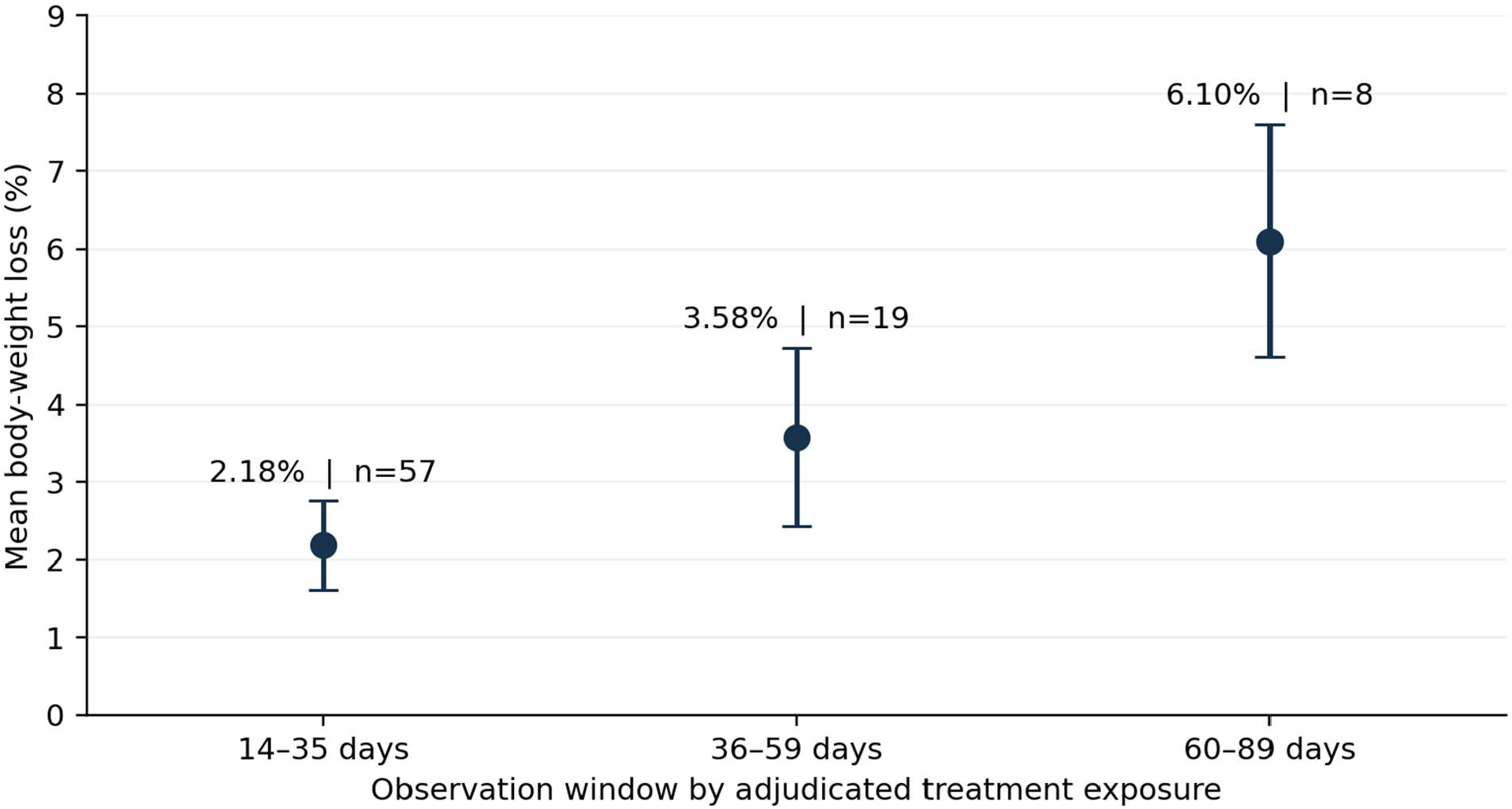
Mean percentage body-weight loss and 95% t-distribution confidence intervals by observation window. Labels give patients contributing one selected measurement per window: 84 patient-window records from 60 distinct patients. All 93 eligible measurements, including nine additional same-window measurements, enter the mixed model. Window populations overlap and are not independent groups.

**Figure 3.**
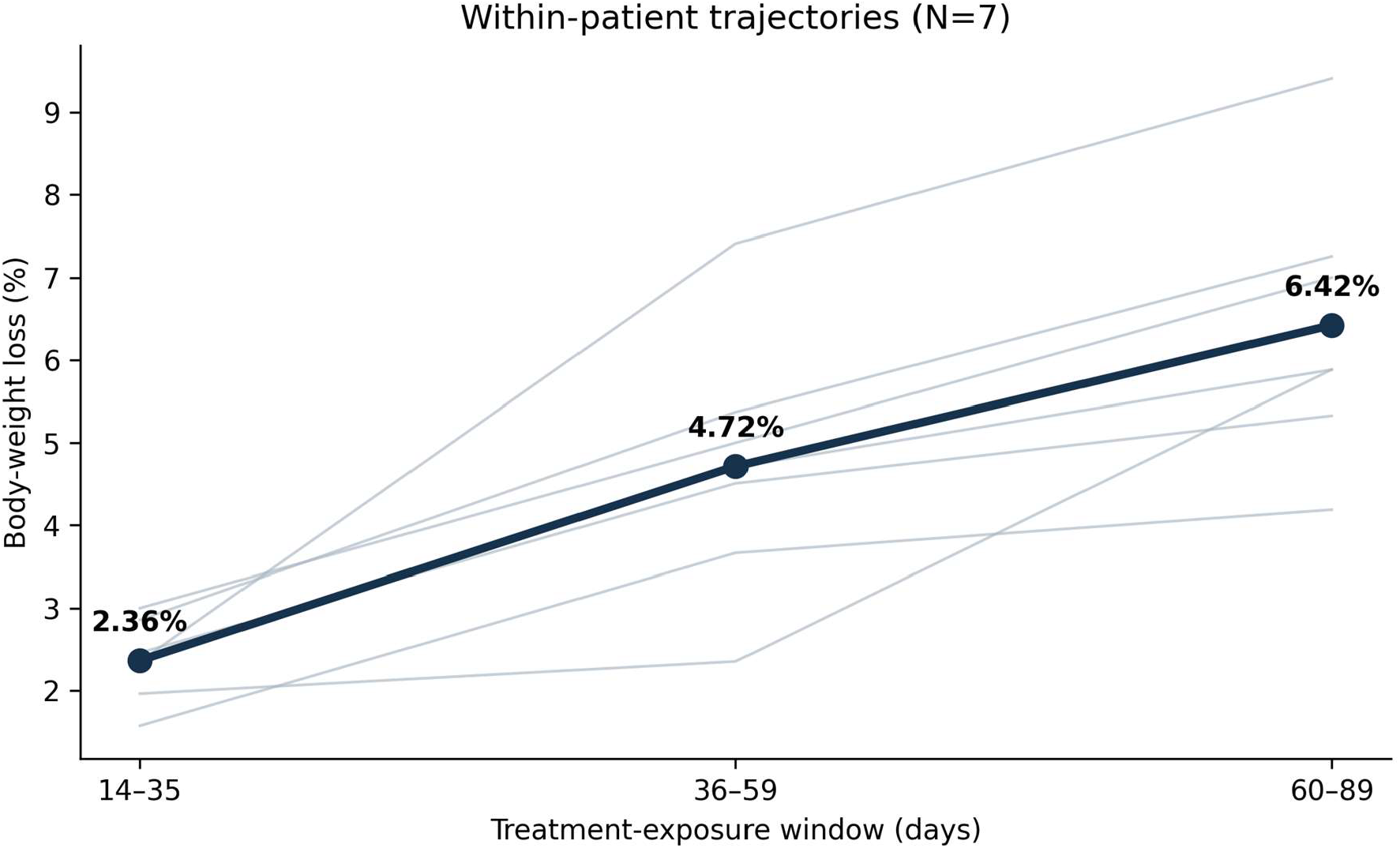
Within-patient trajectories for the seven patients observed in all three windows. They are a subset of the 16 patients observed in both the 14–35- and 36–59-day windows. Thin lines denote individual trajectories; the heavy line denotes the mean. This selected subset is descriptive and does not establish absence of attrition bias.

### Within-patient and longitudinal analyses

Sixteen patients had selected measurements in both the 14–35- and 36–59-day windows. Their mean losses were 2.20% and 4.10%, respectively: an additional 1.91 percentage points (95% CI 1.17–2.64; paired t-test, nominal P<0.001). Seven of these 16 patients also had measurements in the 60–89-day window. For this nested seven-patient subset, mean losses were 2.36%, 4.72%, and 6.42% across the three windows. These paired descriptions do not remove selection bias among patients who continued follow-up.

The adjusted random-intercept model using all 93 measurements from 60 patients estimated 2.16 percentage points greater weight loss per 30 days of elapsed treatment exposure (95% CI 1.69–2.63; nominal P<0.001). This describes a time association conditional on observed follow-up and model assumptions, not a causal treatment effect or randomized dose comparison.

### Initiation at 1 mg/week

Starting dose was taken from the prescription-exposure adjudication table, not from the first qualifying follow-up. Fifty-three of 60 patients (88.3%) initiated at 1 mg/week and seven (11.7%) at 2 mg/week.

Among 1-mg initiators, mean loss was 2.26% (95% CI 1.63–2.90; n=51) at 14–35 days, 3.86% (95% CI 2.83–4.90; n=18) at 36–59 days, and 6.10% (95% CI 4.60–7.60; n=8) at 60–89 days. Some titrated to 2 mg/week; these are initiation-dose groups, not groups maintained at 1 mg/week.

### Patient-reported tolerability

The first eligible frozen follow-up measurement, before window-level selection, was matched to the same patient and calendar date in the intake/refill export. Side-effect and overall-feeling responses were available for 58 of 60 patients; two could not be linked in that export and were treated as missing, not symptom-free. Eleven of 58 (19.0%) reported at least one side effect and 57 of 58 (98.3%) rated overall feeling good or excellent (Table 3). These routine reports are not systematic adverse-event surveillance. Responses were missing/unmatched for 2/60 patients. Symptoms are not mutually exclusive. A zero vomiting report at one check-in is not evidence of zero risk.

**Table 3.** Patient-reported outcomes at the first eligible frozen follow-up.

| Patient-reported outcome | n/N | % |
| --- | --- | --- |
| Any reported side effect | 11/58 | 19.0 |
| Nausea | 7/58 | 12.1 |
| Constipation | 6/58 | 10.3 |
| Diarrhea | 2/58 | 3.4 |
| Vomiting | 0/58 | 0.0 |
| Overall feeling good or excellent | 57/58 | 98.3 |

### Sensitivity analyses

Selecting the first instead of the latest measurement per window yielded mean losses of 2.03%, 3.52%, and 6.21%, respectively (n=57, 19, and 8). In the additional three-day delayed-start scenario, the means were 2.43%, 3.83%, and 6.12% (n=43, 20, and 5); in the seven-day scenario, they were 2.78%, 4.07%, and 6.18% (n=36, 18, and 4). These scenarios retained 76 measurements from 47 patients and 64 from 42, respectively. Changing start assumptions reclassified windows and changed who remained eligible.

## Discussion

Among evaluable adults documented as new starts, mean weight loss during treatment at 1–2 mg/week was 2.2% in the 14–35-day observation window at a median 21 days and 3.6% in the 36–59-day window at a median 48 days. The eight patients with 60–89-day observations had mean loss of 6.1% at a median 65.5 days. The increasing pattern also appeared in selected paired observations and the repeated-measures model, but all findings remain hypothesis-generating.

The study addresses an evidence gap distinct from pivotal tirzepatide trials. SURMOUNT-1 evaluated maintenance doses of 5, 10, and 15 mg/week after labeled initiation and demonstrated substantial 72-week weight reduction.^1^ Our findings are not directly comparable because follow-up was much shorter, treatment was compounded, outcome capture was observational, and no control group was available. The results therefore should not be used to claim that lower doses are equivalent or superior to approved regimens.

Source validation addressed identifiable weight, route, and treatment-timing inconsistencies. Using administrative approval as treatment start would have assigned more elapsed exposure time than the adjudicated estimate. The analysis describes weight change while documented treatment remained at 1– 2 mg/week; it does not attribute post-escalation outcomes to that regimen. However, review followed inspection of provisional outcomes, so independent replication and prospectively specified validation rules remain important.

Important limitations remain. There was no comparator, so medication effects cannot be separated from dietary or behavioral changes, regression to the mean, and other confounding. Weights were self-reported. Inclusion and later observations required documented follow-up; response or tolerability may influence return, and the 60-patient analysis does not represent all 220 otherwise eligible new starts.

Both the 36–59-day (n=19) and 60–89-day (n=8) estimates are based on small selected subsets; the 95% mean-loss intervals span 2.43–4.72% and 4.60–7.60%, respectively. Precision depends on sample size and variability, not a universal cutoff, and no ≥90-day outcome was available. Estimated start dates do not verify adherence. Delayed-start sensitivities change the analyzed population and cannot eliminate timing error, missingness bias, or confounding. The observed 1-versus-2-mg groups are too small and nonrandomized to support a dose-response comparison. Routine symptom capture, the predominantly female cohort, compounded tirzepatide with vitamin B12, and commercial author affiliations further limit safety assessment and generalizability.

Prospective studies should prespecify measured weights, verified administration dates, standardized adverse-event capture, treatment adherence, concomitant therapies, and a clinically appropriate comparator. Randomization or carefully designed comparative effectiveness analyses are needed before lower-dose initiation strategies can be recommended.

## Conclusions

Weight loss was observed within the 14–35-, 36–59-, and 60–89-day observation windows among evaluable adults initiating compounded injectable tirzepatide at 1–2 mg/week. These hypothesis-generating observations support prospective comparative study, not conclusions about causality, equivalent dosing, or long-term safety.

## Declarations

### Data Availability

Individual patient-level data are private and cannot be shared publicly. De-identified data and the analysis code can be provided upon reasonable request to the corresponding author.

### Ethics Approval

This study is a secondary analysis of de-identified data collected during routine clinical care at PlexusDx. As the analysis involves no direct patient contact, no intervention, and uses only data de-identified per HIPAA Safe Harbor [45 CFR 164.514(b)(2)], the PlexusDx Research Committee determined it does not constitute human subjects research as defined by 45 CFR 46.102. Formal IRB review was therefore not required.

### Consent for Publication

Not applicable. The manuscript reports aggregate results and unlabeled individual trajectories and contains no directly identifying patient information.

### Author Contributions

Jay Hastings: Conceptualization, Methodology, Resources, Supervision, Project administration, Writing – original draft, Writing – review & editing. Jayden Lee, PharmD, EMBA: Clinical interpretation, Methodology, Validation, Writing – review & editing. Both authors approved the final manuscript and accept responsibility for the work.

### Data and Code Availability

Individual patient-level data are private and cannot be shared publicly. De-identified data and the analysis code can be provided upon reasonable request to the corresponding author.

## Acknowledgments

Generative AI tools, including OpenAI ChatGPT and Anthropic Claude, assisted with manuscript drafting, code preparation, and revision reconciliation. The authors reviewed and verified the analyses and final wording and take responsibility for the accuracy, integrity, and originality of the work.

